# Development and validation of risk scores for all-cause mortality for the purposes of a smartphone-based ‘general health score’ application: a prospective cohort study using the UK Biobank

**DOI:** 10.1101/2020.11.23.20229161

**Authors:** Ashley K. Clift, Erwann Le Lannou, Christian P. Tighe, Sachin S. Shah, Matthew Beatty, Arsi Hyvärinen, Stephen J. Lane, Tamir Strauss, Devin D. Dunn, Jiahe Lu, Mert Aral, Dan Vahdat, Sonia Ponzo, David Plans

**Affiliations:** Department of Surgery and Cancer, Imperial College London, United Kingdom; Huma Therapeutics, Millbank Tower, London, United Kingdom; SITE Department, University of Exeter, Exeter, United Kingdom; Department of Experimental Psychology, University of Oxford, Oxford, United Kingdom

## Abstract

**Background:** Even though established links exist between individuals behaviours and potentially adverse health outcomes, to date either univariate, simpler models or multivariate, yet difficult to employ ones, have been developed. Such models are unlikely to be successful at capturing the wider determinants of health in the broader population. Hence, there is a need for a multidimensional, yet widely employable and accessible, way to obtain a comprehensive health metric.

**Objective:** To develop and validate a novel, easily interpretable points-based health score (“C-Score”) derived from metrics measurable using smartphone components, and iterations thereof that utilise statistical modelling and machine learning approaches.

**Methods:** Comprehensive literature review to identify suitable predictor variables for inclusion in a first iteration points-based model. This was followed by a prospective cohort study in a UK Biobank population for the purposes of validating the C-Score, and developing and comparatively validating variations of the score using statistical/machine learning models to assess the balance between expediency and ease of interpretability versus model complexity. Primary and secondary outcome measures: Discrimination of a points-based score for all-cause mortality within 10 years (Harrell’s c-statistic). Discrimination and calibration of Cox proportional hazards models and machine learning models that incorporate C-Score values (or raw data inputs) and other predictors to predict risk of all-cause mortality within 10 years.

**Results:** The cohort comprised 420,560 individuals. During a cohort follow-up of 4,526,452 person-years, there were 16,188 deaths from any cause (3.85%). The points-based model had good discrimination (c-statistic = 0.66). There was a 31% relative reduction in risk of all-cause mortality per decile of increasing C-Score (hazard ratio: 0.69, 95% CI: 0.663 to 0.675). A Cox model integrating age and C-Score had improved discrimination (8% percentage points, c-statistic = 0.74) and good calibration. Machine learning approaches did not offer improved discrimination over statistical modelling.

**Conclusions:** The novel health metric (‘C-Score’) has good predictive capabilities for all-cause mortality within 10 years. Embedding C-Score within a smartphone application may represent a useful tool for democratised, individualised health risk prediction. A simple Cox model using C-Score and age optimally balances parsimony and accuracy of risk predictions and could be used to produce absolute risk estimations for application users.

## Introduction

Despite the empirical establishment of strong relationships between given behaviours, lifestyle factors and the development of preventable diseases, individuals may struggle to tangibly conceptualise how their day-to-day behaviour impacts their long-term health outcomes. A number of ‘health metrics’ have been developed to quantify general health at a given point in time and estimate risk of negative future outcomes, however few of these tools are accessible, interpretable, actionable and easy to calculate. The use of univariate measures whilst easily calculable and interpretable may incompletely capture the wider determinants of health, such as psychological wellbeing, for example.

Body Mass Index (BMI) is often used as a quick means of estimating an individual’s relative adiposity and infer the relative likelihood of adverse adiposity-related outcomes [1,2]. Despite relative ease of calculation, BMI has numerous oft promulgated limitations, including issues with scalability (two people with same body proportions but different heights will have divergent BMI), its ignorance of variation in physical characteristics due to age, sex or ethnicity, [3,4] its inability to discern between muscle and fat, and a variable strength of relationship to health outcomes [5].

Multivariable risk prediction models can be easily developed using statistical modelling [6,7] or machine learning [8,9] approaches on appropriate datasets and lend themselves to supporting decision making across the manifold aspects of health and disease management or prevention. Indeed, there are a multitude of models published that seek to predict all-cause mortality [10,11]. All-cause mortality is an easily understandable risk trajectory into which the natural histories of many preventable diseases converge, and can be manipulated by behavioural changes. Therefore, it represents an attractive target for a general health metric or predictive model. There are however hurdles to the widespread use of such predictive models [12]. Validation of models in the datasets that they were derived from (internal validation) and also an assessment of their ability to ‘generalise’ to independent datasets, preferably in different populations (external validation) must be achieved prior to widespread use [13,14]. However, even if validated, models tend to remain the preserve of clinicians and may incorporate mathematical analysis of data points that require invasive testing (e.g. blood tests), may be non-modifiable by ‘users’ (e.g. childhood exposures) or are not easily accessible (e.g. deprivation index).

Therefore, an unmet need in public health is the presence of validated health metrics based on models that are not only strongly predictive of outcomes, but also accessible, have an understandable/interpretable output, and are parsimonious. Furthermore, should causal mechanisms be clearly established and the metrics validated as ‘causal prediction models’, the focussed use of modifiable predictor variables could help demonstrate actionable insights to guide beneficial lifestyle changes. With the ability of smartphones to utilise inbuilt hardware to capture multimodal data relevant to physiological status, we believe that a smartphone application integrating product design, technological and risk modelling principles could present a novel conduit for risk prediction models focussing on well-established risk factors to enable members of the general public to engage with their health.

Here, the authors describe the development of a novel multivariable health metric, hereon named “C-Score”, which seeks to mathematically integrate parameters that can be measured digitally, are almost all modifiable, and are relevant to various domains of health. Three formats of risk score or model are developed: a simple and easily interpretable 0-100 points-based score developed by summation of published literature regarding multiple variables across multiple geographies; statistical modelling using Cox proportional hazards methods analysing C-Score with other predictors such as age, and also machine learning models for the same. The first is validated, and the latter are both developed and validated using the UK Biobank [15] data.

## Methods

This study sought to develop and validate forms of novel risk models for the purposes of a general health metric suitable for embedding within a smartphone application. Given the convergence of multiple key risk factors on the risk of all-cause mortality as well as morbidity and mortality from leading non-communicable diseases, the target endpoint chosen for this metric was all-cause mortality. The study was planned and conducted in accordance with TRIPOD guidelines [26].

### Candidate explanatory variables for models

A comprehensive literature review was undertaken using PubMed for candidate predictor variables for all-cause mortality. Search terms included “all cause mortality”, “death”, “mortality prediction” and “risk model”. In addition, pre-posited candidate variables were searched alongside “all cause mortality”, such as “smoking AND all-cause mortality, or “resting heart rate AND all-cause mortality”.

The candidate variables that were identified from the literature review, which was led by clinical and epidemiological acumen regarding biological plausibility, were considered by the authorship panel in terms of their evidence base, but also the degree to which they are modifiable, their ability to measured using inbuilt capabilities of commonly available smartphones, and lastly their contributions to engaging user design perspectives. As the intention was to develop an interpretable ‘general health metric’ that will be relevant to multiple morbidities and not only mortality, candidate variables were reviewed in terms of overlap with leading causes of morbidity and mortality.

Ultimately, eight predictor variables were selected: age, cigarette smoking, alcohol intake, self-rated health, resting heart rate, sleep, cognition (reaction time) and anthropometrics (waist to height ratio).

### Development of a points-based score (C-Score)

The first risk index (“C-Score”) attempted to formulate an easy-to-interpret continuous score that used published evidence from multiple countries focussed chiefly on modifiable factors, as opposed to developing a model using a single large database from one geography. This approach sought to utilise published hazard ratios or regression coefficients to weight individual parameters, as has been done elsewhere [16].

Identified studies as per above search criteria were reviewed by the authorship panel for cohort size, length of follow-up, the robustness of statistical analyseis, and whether or not hazard ratios for all-cause mortality were reported (also if these were adjusted for age, gender and/or other confounders).

We opted to include all important variables other than age in the first iteration of the points-based model to ascertain the power of purely dynamic/modifiable characteristics in a risk index.

Hazard ratios were extracted from the studies deemed to be of the highest quality by the authorship panel. These were used as relative ‘weightings’ for a points-based score. The optimal value for each input was set as 0 (lowest risk), with increasing numbers of points assigned for greater diversions away from optimal risk level – these points reflected the literature-extracted hazard ratios. The raw sum of maximal hazard ratios was approximately 25; therefore, the values for all increments values of hazard ratio were quadrupled to make a total sum of 100.

The points-based score functions in a penalising fashion, that is, users ‘start’ with 100 points: for each health domain, they can sequentially either lose no points (if they optimise that data input) or lose points in accordance with the HR associated with that level of exposure. For example, users will lose no points for being a never-smoker but will lose more points if they smoke over 20 cigarettes per day versus a smoker of 10 per day. As such, the output from the score is a continuous variable; a number between 0 and 100, where 100 is optimal. Table 1 demonstrates the maximum penalty for each sub-domain, as informed by extracted hazard ratio data [11,17–22].

**Table 1.** The range of points allocated to ‘users’ per data input to C-Score. C-Score is calculated by subtracting total number of points from 100, to yield a 0-100 score wherein 100 is optimal.

| **C-Score input metric and ‘domain’** | **Range of points allocated** |
| --- | --- |
| Resting heart rate (beats per minute)<br>(Cardiovascular fitness) | 0 – 7.83 |
| Average hours of sleep per night<br>(Sleep habits) | 0 – 10.26 |
| Waist-to-height ratio<br>(Adiposity) | 0 – 10.8 |
| Self-rated health (ordinal scale: Excellent, Good, Fair, Poor)<br>(Surrogate for existing co-morbidity or perception of ill-health) | 0 – 31.32 |
| Cigarette smoking (status and cigarettes per day)<br>(Tobacco exposure, including past smoking) | 0 – 12.96 |
| Alcohol consumption (units/week)<br>(Alcohol intake) | 0 – 19.44 |
| Reaction time (milliseconds)<br>(Neurocognition) | 0 – 6.75 |

As the C-Score does not output a percentage risk prediction, it was only assessed for discrimination in predicting all-cause mortality within 10 years. As is evident below, percentage absolute risk assessments are possible if the C-Score is incorporated as a variable in statistical modelling approaches.

### Data collection methods and materials

The raw data contributing to the C-Score calculation is obtained by means of an ad-hoc smartphone application (Figure 1). Data inputs for the score included manual entry (e.g. age, gender, alcohol and tobacco consumption), image analyses (e.g. waist-to-height ratio) and use of phone camera technology (e.g. RHR). Waist-to-height ratio was calculated using camera-based anthropometric measurements (BVI), which produced the following outputs: waist circumference, hip circumference, body fat percentage and total body volume. Finally, RHR was collected via a camera-driven photoplethysmogram sensor, which is able to detect heartbeats when participants position their finger on the camera.

**Figure 1.**
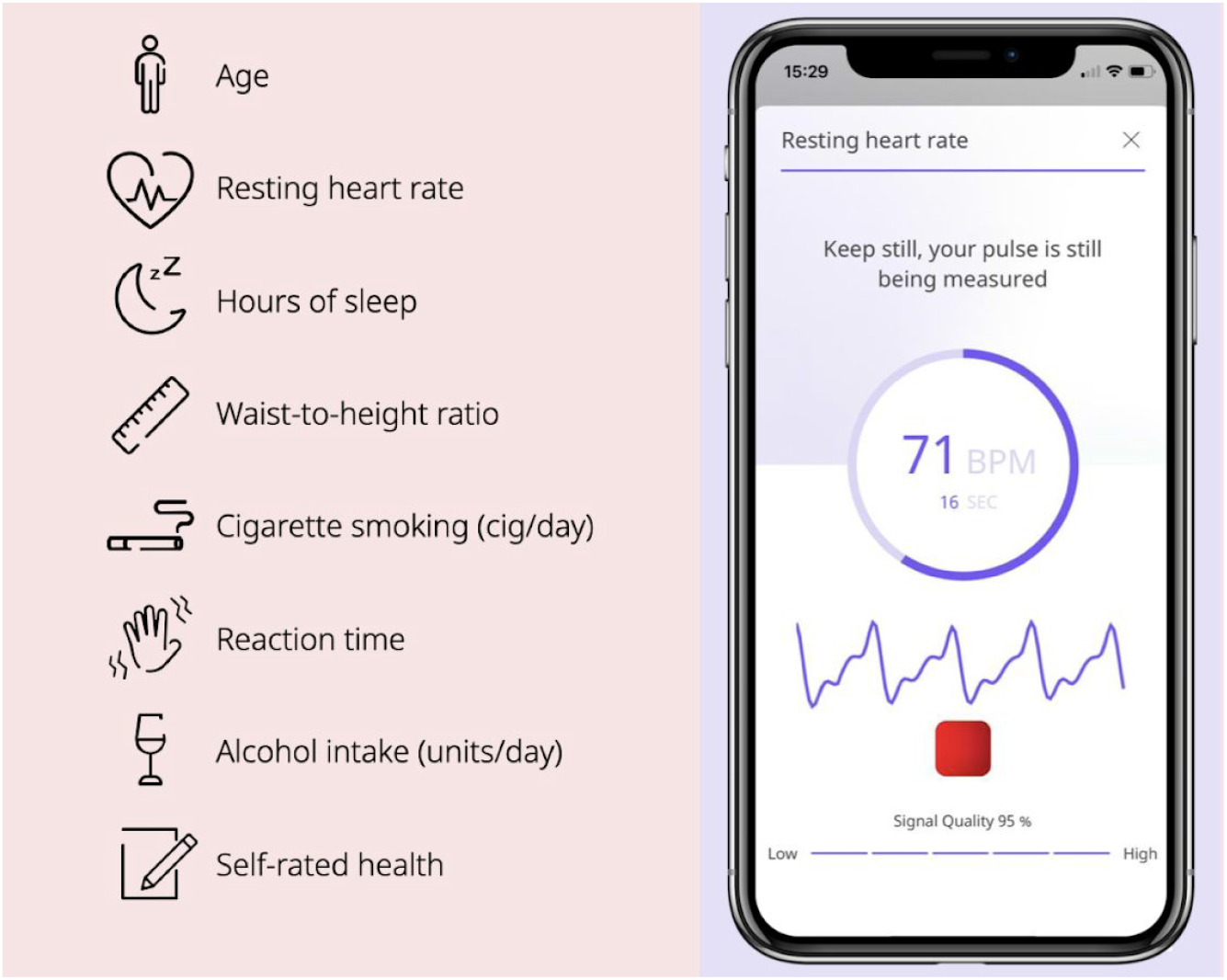
Screenshots of the C-Score mobile application. A) List of data contributing to the C-Score; B) RHR measurement screen and C) body scan screen (for waist-to-height ratio).

### Study population

The individual-level data of the UK Biobank was utilised as a study population for the validation of the points-based model, and also the development and validation of risk models for all-cause mortality that incorporate C-Score inputs (as more complex variants of the initial points-based score). There were no specific eligibility criteria for inclusion in our analyses; the entirety of the available dataset was used for our analyses. Briefly, the UK Biobank represents a prospective cohort study wherein over 500,000 individuals aged 40-69 were recruited between 2006-2010 and followed up thereafter [15]. Participants underwent a robust assessment centre at baseline where multiple questionnaires were completed, anthropometric measures were taken, and also blood/saliva samples were obtained. Aside from robust phenotyping, participants have also been genotyped, 100,000 are in the process of undergoing whole body imaging, and 20,000 have completed repeat assessments. Participants data are linked to multiple other electronic databases, enabling ascertainment of among others, data regarding date of death and cause of death. The UK Biobank is an ‘open’ resource accessible to any researchers approved as *bona fide* by the Biobank Access Management Team.

### Development of de novo models

Whereas C-Score was validated in terms of discrimination using the UK Biobank, three further versions of models were both generated and validated using data in the UK Biobank, referred to as Models 2 to 5. As the continuous ‘health score’ does not predict percentage risks, we used Cox regression to form variants of models that can output such a risk prediction and assess their discrimination and calibration. This opens the possibility of having a user-facing score, with scope for generating individualised percentage-style risk estimates for multiple outcomes of interest, such as all-cause mortality.

Model 2 integrated C-Score and age, whereas Model 3 integrated the raw values for all C-Score inputs and also age to assess the amount of performance sacrificed by a predetermined weighting system. Model 4 sought to develop ‘maximally complex’ statistical models with interactions to identify maximal-attainable predictive accuracy and also included sex and ethnicity, again to assess the balance between predictive power and expediency of a simple, interpretable score or simple model. These Cox models were developed to predict the risk of death within 10 years of follow-up as a complete case analysis. As the intended smartphone application would require completion of all data fields to complete the health score, a complete case analysis of UK Biobank data offered a form of evaluation that most closely aligned to the intended use of the models. The baseline data values (i.e. obtained from the assessment centre) were used to calculate C-Scores and also participant baseline age for the development of the Cox models. Individual follow-up was defined as time elapsed from initial assessment to either death from any cause or censoring (lost to follow-up or reached the end of study date). End of study date was set as 9^th^ February 2020, which corresponds to the date of data extract download.

### Development of machine learning-based models

The approach taken for Model 5 was the use of supervised machine learning (ML). The problem was specified as a binary classifier, aiming to assign a label representing whether or not the patient dies 10 years post-baseline. Two commonly used supervised machine learning classifiers were chosen, the K-Nearest Neighbour (KNN) Classifier and the Supervised Vector Machine (SVM) Classifier. Both these algorithms were tuned to select the optimal hyper-parameters using 10-fold cross-validation on the training and validation sets before finally being used on a previously unseen 20% of the data to determine the final scores.

As discussed previously, in the UK Biobank, the number of occurrences of the desired outcome is relatively low (less than 5%). In order to deal with this sparsity in the dependent variable, the initial 70:30 train/test split was performed on all the complete cases of the dataset (train n = 319,652 and test n =136,995). However, the actual model was trained by randomly under-sampling the training data (in order to achieve a 50/50 split between the two outcome variables). It is also important to note that these two algorithms are based in feature space, and so the weighting of each feature plays a crucial role in the determination of the classification coefficients. As such, it is important to standardise all the inputs, this was performed by first subtracting the mean and subsequently dividing by the standard deviation for each feature.

We first developed and trained a KNN algorithm to derive a binary label determining the patient’s risk of death in the next ten years. KNNs are a type of classification algorithm based on the premise that similar cases (in feature space) will have similar results. The idea is to classify each new observation based on a metric of ‘nearness’ to all other points and to set its label as the most common label of the k most similar training examples. To use the KNN algorithm two hyper-parameters need to be specified: the value of *K*, i.e. how many training examples will it aggregate to determine the label of the test and the metric for defining ‘nearness’. For both these parameters, we tuned our model using 10-fold cross-validation.

The hyper-parameters used for defining ‘nearness’ are the two most commonly used distance metrics namely the Euclidean distance and the Manhattan distance. The second parameter to be tuned was the value of *K*, values between 5 and 500 were tested. The optimal parameters were determined by maximising the area under the receiver operating characteristic curve (AUROC) using 10-fold cross-validation.

We trained an SVM Classifier to optimally separate in feature space between patients. SVMs are a category of classifiers that aim to determine the hyper-plane that optimally separates the observations into two sets of data points. The intuitive idea behind the SVM algorithm is maximizing the probability of making a correct prediction by determining the boundary that is the furthest from all the observations.

Similar to the previous KNN model, considerations in training have been taken into account and to choose the hyper-parameters. In the case of SVMs, the parameters we have chosen to optimise are, the shape of the separation kernel (linear, polynomial or radial basis function (rbf)), the C regularisation parameter, the degree of the polynomial (only true for polynomial kernels) and the gamma kernel coefficient (for polynomial and rbf kernels). To optimise these parameters, we used 10-fold cross-validation on the training data to maximise the AUROC.

### Statistical analyses and model validation

Continuous variables for descriptive statistics are presented as means and standard deviations. Cox models were developed using the entire available dataset and then underwent internal validation using bootstrapping with 200 iterations (for discrimination and calibration). Models were tested for proportional hazards assumptions (using log-log plots) and inclusion of restricted cubic splines or logarithms for continuous variables.

Discrimination refers to the ability of a prediction model to distinguish between individuals that experience an outcome of interest and those that do not. Suitable metrics include Harrel’s c-statistic which for Cox models is equivalent to the AUROC. A value of 0.5 means that the model is no better than tossing a coin, whereas a value of 1 means perfect prediction.

Calibration refers to the assessment of closeness between predicted and observed risks. This can be assessed by plotting the observed and predicted risk across different levels, such as by tenth of risk. However, ‘binning’ of risk levels is not optimal, and other approaches include linear adaptive spline hazard regression which interpolates across levels of risk [23]. Therefore, we assessed calibration of models using smoothed calibration plots to compare predicted and observed risks, which can also incorporate bootstrapping to correct for model optimism.

Bootstrap optimism-corrected values for the c-statistic were computed, and for Models 2-4, calibration plots were formed. Initial data handling was performed using Stata v16.0 software, with the statistical analyses handled in R statistical software, notably the *rms* package. For Model class 4, algorithms were developed using Python, including the pandas, numpy and sklearn packages; the AUROC is presented.

### Ethical approval

Access to anonymised data for the UK Biobank cohort was granted by the UK Biobank Access Management Team (application number 55668). Ethical approval was granted by the national research ethics committee (REC 16/NW/0274) for the overall UK Biobank cohort.

## Results

### Study population characteristics

In the complete case analysis, there were 420,560 individuals with complete data regarding age at an assessment centre, and all metrics included in ‘C-Score’. There was a maximum follow-up of 13.9 years, and the total follow-up time for the cohort was 4,526,452 person-years. During this, there were 16,188 deaths (3.85% of the cohort).

Demographics for the study cohort were as follows: mean baseline age was 56.58 (SD: 8.07), mean resting heart rate 69.84 (SD: 11.68), mean waist to height ratio was 0.54 (SD: 0.075), mean weekly alcohol intake was 14.34 units (SD 18.84), mean reaction time was 558.03 ms (SD: 117.07), and mean sleep duration was 7.16 hours (SD 1.26). For self-rated health, 68,926 (16.39%) were “excellent”, whereas 245,171 (58.30%), 88,195 (20.97%) and 18,268 (4.34%) were ‘good’, ‘fair’ and ‘poor’, respectively. There were 230,798 men (55.14%), and 188,601 women (44.86%) women. Regarding ethnic background, we grouped these into ‘white’ (397,763: 94.92%), ‘mixed’ (2,480, 0.59%), ‘Asian or Asian British’ (7,631: 1.82%), ‘Black or Black British’ (6,370: 1.52%), ‘Chinese’ (1, 293: 0.31%) or ‘Other’ (3,524: 0.84%).

Regarding calculated C-Scores, the mean score for participants was 77.25 (standard deviation 12.96; minimum 3.34, maximum 100; Figure 2). Figure 3 displays the risk of death within 10 years per C-Score decile.

**Figure 2.**
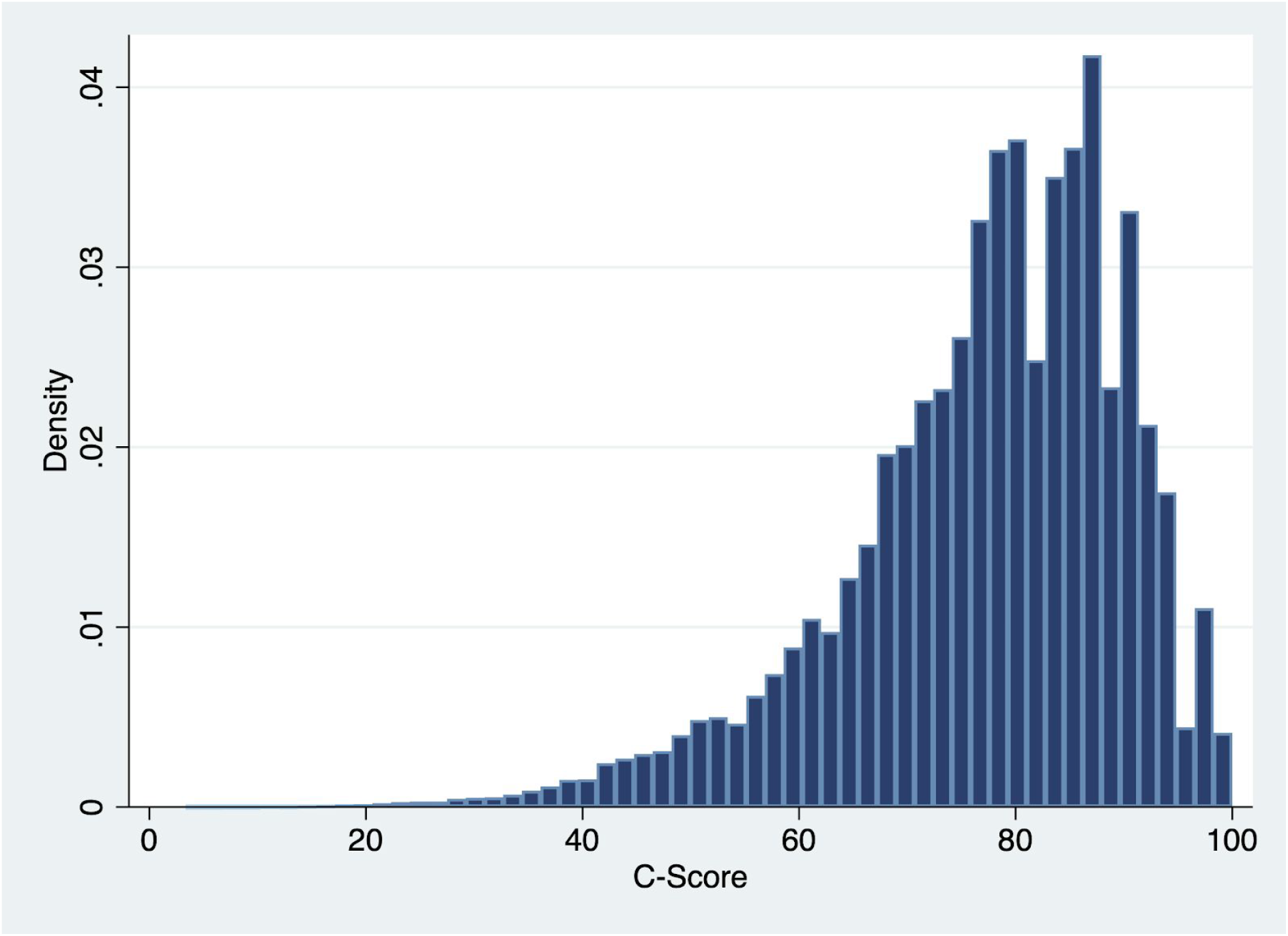
Distribution of C-Score values in the study cohort (n=420,560).

**Figure 3.**
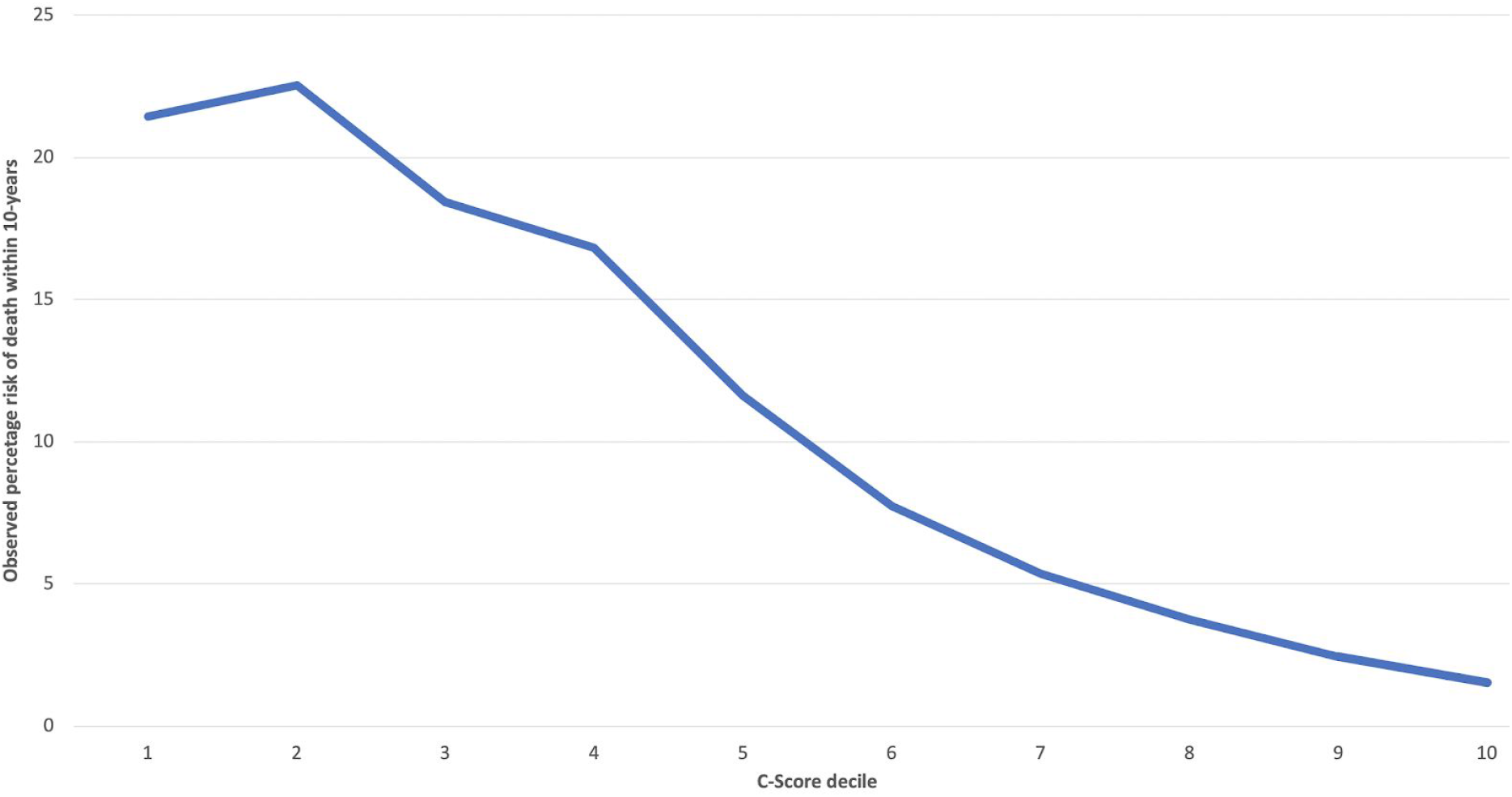
Probability of death within the next 10 years as a function of C-Score decile.

### Discrimination and calibration

Model 1: The hazard ratio for per-unit increase in C-Score was 0.96 (95% CI: 0.960 to 0.961), suggesting a 3% relative risk reduction per unit improvement. When analysed in terms of C-Score decile, i.e. 10-point brackets of C-Score, the hazard ratio was 0.69 (95% CI: 0.663 to 0.675), implying a 31% relative risk reduction of all-cause mortality per decile improvement in C-Score. Regarding discrimination, the c-statistic was 0.66.

Model 2: Inclusion of (log)age and C-Score in a Cox model yielded a c-statistic of 0.74, i.e. an increase in discrimination capability of 8 percentage points. The model appeared well-calibrated (Figure 4). Although age is clearly a major predictor of all-cause mortality, the Cox model demonstrated that on the inclusion of age and C-Score, there were significant roles for both: hazard ratio per year increase in age was 1.09 (95% CI: 1.091 to 1.096), hazard ratio per 10 unit increase in C-Score was 0.67 (95% CI: 0.668 to 0.681). Table 2 demonstrates the coefficients for all Cox models developed.

**Table 2.**
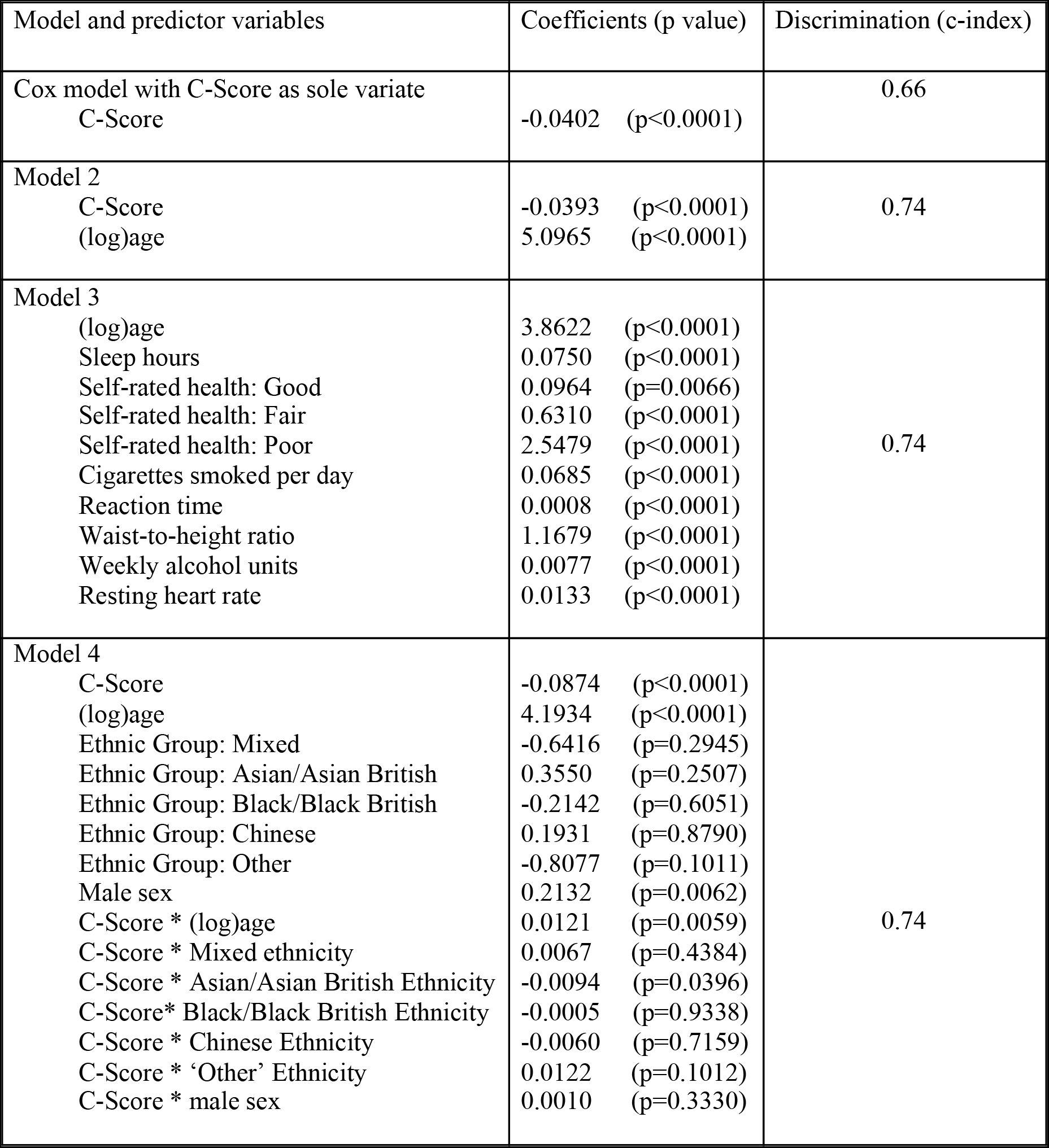
Coefficients from Cox proportional hazards models either examining C-Score alongside confounders/interactions, or raw data inputs (Model 3). Model 4 is presented prior to backwards selection. * denotes interaction term. For reference in terms of discrimination, a Cox model using simply C-Score as the sole variate was developed to demonstrate scope for incremental gains in accuracy.

**Figure 4.**
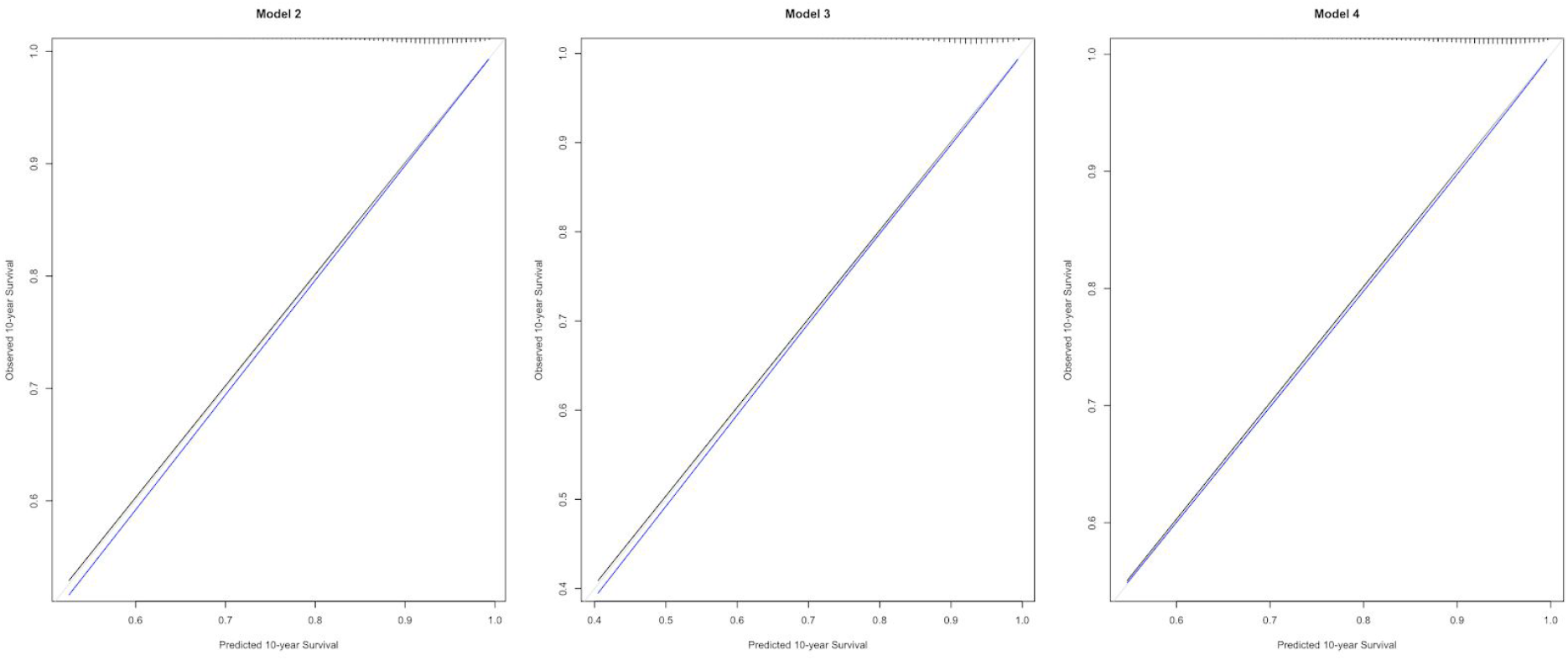
Calibration plots of predicted vs observed probabilities of all-cause mortality within 10 years for Models 2, 3 and 4.

Model 3: Using the raw data inputs rather than a pre-assigned ‘weighting’ plus age yielded a c-statistic of 0.74, therefore no significant improvement in discrimination with a more complex model. The model was also well-calibrated (Figure 4).

Model 4: We also developed a ‘full model’ that included C-Score, age, ethnicity and sex, as well as interactions between C-Score and the latter three. We performed backwards elimination to identify the strongest possible performing model via bootstrapping with 200 iterations; selection was based on the Akaike Information Criterion with a p-value of 0.01. Herein, the final model that retained C-Score, sex, age, an interaction between C-Score and Ethnicity, and an interaction between C-Score and age had an optimism-corrected c-statistic of 0.74, i.e. no improvement with a far more complex model.

Model Class 5: Both the Machine Learning algorithms, KNN and SVM were applied to a test cohort (n_test = 125, 966) in order to predict risk of death in the next 10 years. For the KNN - Following 10 Fold cross-validation for the tuning of the hyper-parameters, we opted to use *K* = 100 and the Manhattan distance metric, the C-index on the test data was 0.72. Similar to the KNN algorithm, we tuned the SVM using 10-fold cross-validation on randomly under-sampled training data. This led us to choose the optimal hyper-parameters as: C = 100, gamma = 0.001, and kernel shape as a radial basis function. This model yielded a C-index of 0.74 but took considerably more hours to train than Model 2.

## Discussion

Risk prediction models have significant potential for assessing the risk of protean events of interest. However, these are limited almost exclusively to clinical use, and widely used/easily accessible health metrics such as BMI have limitations. Extant multivariable prediction models for all-cause mortality are typically poorly accessible to members of the public and risk limited engagement due to perceived non-modifiability of covariates or understanding the mechanisms by which covariates may predict outcomes.. Therefore, for the purposes of this initiative, we opted to migrate away from univariate assessments towards a novel, multivariable health metric focussed on characteristics that span multiple domains of health, is accessible and can be used by anybody with a smartphone application. Our results demonstrate that the value of an easily interpretable points-based score to infer all-cause mortality risk, and mandate consideration of this smartphone-based health index in the prediction of multiple other diseases or conditions. Our results also suggest that a simple Cox model including C-Score and age may provide accurate absolute risk predictions for public health initiatives, such as promoting public understanding of individual health risk, or raising awareness of the effects of behaviours on health. Lastly, they are interesting regarding the power of statistical modelling approaches relative to machine learning approaches using the same data.

All-cause mortality was selected as a first endpoint for validation purposes given its ease of comprehension and its close links to multiple modifiable behaviours and/or it oft being a consequence of preventable disease. This is an end-point that has been robustly examined in the UK Biobank by two other key studies. A study by Weng, et al. [10] utilised the UK Biobank to derive epidemiological (Cox) and machine learning models (random forests and neural networks) to predict premature mortality using a pre-selected panel of 60 candidate baseline predictor variables encompassing aspects such as socio-demographics, educational attainment, behaviour, nutritional intake, lifestyle factors, medication use and clinical history. In standard Cox modelling, the final included variables were gender, log age, educational qualifications, ethnicity, previous diagnoses of cancer, coronary heart disease, type 2 diabetes and COPD, smoking history, blood pressure, Townsend deprivation index and BMI. Variables included in random forests modelling included BMI, FEV1, waist circumference, blood pressure parameters, skin tone and age. On identifying the optimal neural network parameters identified by grid-search from 10-fold cross-validation, top risk factor variables included smoking status, age, prior cancer diagnosis, prescription of digoxin, residential air pollution and Townsend deprivation index. The discrimination of the fully adjusted Cox model, random forests and neural network were 0.751 (95% CI: 0.748-0.767), 0.783 (95% CI: 0.776-0.791) and 0.79 (95% CI: 0.783-0.797), respectively [10]. Whilst these AUROC values are significantly but marginally higher than those reported with our intended app-based model, they included variables that are for the most part non-modifiable and do not offer clear scope for use by members of the public to not only compute their risk, but also be able to act on various components to reduce risk.

Ganna and Ingelsson used the UK Biobank cohort to identify predictors of 5-year all-cause and six cause-specific mortality categories from 655 measurements of demographic, the results of which were interestingly packaged as part of an interactive website named UBBLE [11]. Ultimately, for all-cause mortality within 5-years, 13 predictors for men and 11 for women in a Cox model achieved discrimination of 0.8 (95% CI 0.77-0.83) and 0.79 (95% CI: 0.76-0.83). Again, although these models attained a significantly but slightly higher discrimination than our model, the majority of parameters included are minimally modifiable (in retrospect, number of children given birth to), difficult to explain their effect on mortality (number of people in the household, numbers of cars or vans owned/used by household, relationships of people lived with) and emphasise existing health conditions (known diabetic, previous cancer) [20]. Whilst our study did not validate the C-Score as a ‘causal prediction model’ where coefficients have a direct causal interpretation, such further work is underway, and the inclusion of modifiable factors known to have causal implications on health outcomes is encouraging in this regard.

In the era of ‘big data’, a resurgence in the popularity of artificial intelligence (AI) and more specifically ML has been seen across a wide array of fields including healthcare. These novel methodologies have led to some notable results in prediction and diagnostics and so have become a commonly examined tool in medical research. It is, however, important to note that ML techniques do not always lead to better results than ‘classical’ statistical methods. Indeed, we observed comparatively similar or even lower results using two very popular and widely used algorithms, namely KNN and SVM, than using a traditional epidemiological/statistical modelling strategy. Furthermore, due to the inherent complexity associated with ML algorithms, these took far longer to train and required greater computational power. ML methodologies rely on the artificial generation of knowledge using machine-guided computational methods instead of human-guided data analysis in order to find a best fit in the data. This has some very strong use cases, especially when dealing with wide and complex datasets with multifactorial causation, complex and potentially non-intuitive interactions. However, in this study, we show that ML is not always the answer and that initial development of an algorithm with few metrics and careful consideration of the input by those with scientific/clinical acumen can yield higher results.

Our work has some strengths and also limitations. Strengths include the use of the UK Biobank which provides a contemporary, richly phenotyped cohort with linkages to national registries, which minimises loss to follow-up, prospectively evaluates risk factors and enables accurate ascertainment of outcomes of interest. Another strength is the cognisance of target users of the app that the model is intended for, which drove us to focus on modifiable risk variables where possible - we were content with sacrificing a small percentage of discriminatory capability without needing to ‘penalise’ intended users for having pre-existing conditions, a certain educational level, or living within air pollution.

Possible limitations of our work include the use of a complete case analysis which may have introduced bias, and the use of ‘human intelligence’ to prune the possible list of candidate predictor variables which limited the scope for ML to perform optimally. As the overwhelming majority of missing data for the included variable were due to participants “not knowing” the answer or refusing to answer, we considered this to replicate the target end-situation where people will be using the health index or model in an app. We mitigated bias to the best of our abilities throughout the rest of methodology for modelling where possible - for example, we used the whole dataset for Cox modelling and bootstrapping for validation, rather than randomly splitting the data into development and validation set which is inefficient and inadvisable [7,12]. The fact that machine learning methods did not deliver significant improvements in discrimination is not a formal comparative assessment of statistics versus machine learning - indeed, machine learning is likely best ‘reserved’ for situations other than trying to optimise the weighting of a small number of variables that humans have pre-selected. There is also the limitation of selection bias with regards to the participants of the UK Biobank, as they tend to be slightly ‘healthier’ than the general population at large [24,25]. Use of the C-Score outside the UK population and in different age groups should follow validation in appropriate local datasets, work on which is underway.

In conclusion, we believe that the ‘general health metric’ reported here not only compares well to other work despite using fewer variables, but offers several advantages from a population-use perspective as it offers a holistic review of multiple aspects of health and focuses for the most part on modifiable characteristics that could in time be targets for risk-reducing interventions pending further model evaluation. Our proclivity was not to produce the most powerfully predictive models possible using a prospective data set, but rather develop and validate models that are rational, understandable and could be engaging within a smartphone app. Given the strong association of many of the included variables on other diseases (and not just all-cause mortality), we believe that a points-based score may be powerful in making inferences regarding current and future health in terms of individual conditions. Even more powerful could be simple statistical models incorporating C-Score and age for each clinical endpoint of interest. Further work is already underway within our group to assess the capabilities of C-Score and variations thereof across a panel of conditions of interest, as is the embedding of this score system within a mobile application.

## Data Availability

The UK Biobank cohort data is available to researchers as approved by the Biobank Access Management Team. Due to commercial sensitivity, we have not presented the complete raw weighting system for deriving the C-Score here: this could be made available by Huma to academic partners seeking to collaborate to externally validate C-Score models in other datasets. The R/Python code used by the investigators for Cox modelling/ML modelling can be provided on request to the authors.

## Funding statement

Funding for the purposes of this project was provided by a contract between Chelsea Digital Ventures and Huma Therapeutics (previously known as Medopad). Funders had no role in the data acquisition, data analysis or the write-up of this manuscript.

## Competing interest statement

AKC is a previous consultant for Huma. DP, SP, ELL, CPT, SSS, MB, AH, TS, DD, JL, MA, DV, and SL are employees of Huma Therapeutics.

## Contributions

AKC: Design of study, acquisition and analysis of data, manuscript writing

ELL: Design of study, analysis of data, manuscript writing

CT: Conception of study, study design, manuscript writing and critical review

AH: Conception of study, study design, critical revision of manuscript

All other authors: conception of study and design, critical revision of manuscript. All authors have approved the final version of the manuscript submitted. All authors agree to be accountable for all aspects of the work in ensuring that questions related to the accuracy or integrity of any part of the work are appropriately investigated and resolved.

